# Characterization of Natural Killer Cell Subpopulations in the Blood of Single Ventricle/Hypoplastic Left Heart Syndrome Patients Using Single-Cell RNA Sequencing

**DOI:** 10.1101/2024.08.18.24312189

**Authors:** Hui-Qi Qu, Kushagra Goel, Kayleigh Ostberg, Diana J Slater, Fengxiang Wang, James Snyder, Cuiping Hou, John J Connolly, Michael March, Joseph T Glessner, Charlly Kao, Hakon Hakonarson

## Abstract

We utilized single-cell RNA sequencing (scRNA-seq) to examine peripheral blood mononuclear cells (PBMCs) from patients with Single Ventricle/Hypoplastic Left Heart Syndrome (SV/HLHS), and demonstrated a more pronounced correlation between gene expression in Natural Killer (NK) cells and SV/HLHS compared to other PBMC cell types. Our scRNA-seq analysis of NK cells in this study identified two distinct clusters with gene expression patterns linked to immune responsiveness and adaptation to stress. While this finding underscores the heterogeneity of NK cells, it provides new insights into fine-tuning of immune modulation that could prevent complications in SV/HLHS. Specifically, our study suggests that while NK cells in SV/HLHS are adapting to support survival in a challenging physiological environment, these adaptations may compromise their ability to manage additional stresses such as infections and inflammation.

## Introduction

Natural Killer (NK) cells are essential in the body’s innate immune response(Colucci et al. 2003). In our study of single-cell RNA sequencing(scRNA-seq) from peripheral blood mononuclear cells (PBMCs) of patients with Single Ventricle/Hypoplastic Left Heart Syndrome(SV/HLHS), we found that gene expression in NK cells is more closely correlated with SV/HLHS than in other cell types by weighted gene co-expression network analysis (WGCNA)(Qu et al.). In congenital heart diseases (CHD) including SV/HLHS, NK cells play a crucial role in managing infections and mediating complications related to CHD(Wienecke et al. 2022). In our previous study, we identified 1600 genes that showed differential expression (DE) in NK cells of SV/HLHS patients(Qu et al.). Our current investigation suggests that NK cells in SV/HLHS patients are heterogeneous and potentially receptive of different types of immune modulation. We hypothesized that the SV/HLHS-related DE genes might be affected differently across specific NK cell subpopulations, instead of uniformly across all NK cells. To test this hypothesis, we delved into the single cell transcriptome of NK cells in greater detail.

## Methods

### scRNA-seq of PBMCs

This study received approval from the Institutional Review Board at the Children’s Hospital of Philadelphia (CHOP). PBMCs from three de-identified children (2 males and 1 female) with SV/HLHS were compared to those from three healthy controls (2 males and 1 female). Blood samples were collected in EDTA-coated tubes and promptly processed at the Center for Applied Genomics (CAG) at CHOP. PBMCs were isolated using Ficoll density gradient centrifugation. scRNA-seq was conducted using the 10X Chromium Single Cell Gene Expression assay (10x Genomics, Single Cell 3’ v3)(Qu et al. 2023). Sequencing was performed on the Illumina HiSeq2500 SBS v4 platform. The resulting data from the Chromium scRNA-seq were processed with the Cell Ranger 7.1.0 software suite (10x Genomics), with sequencing reads aligned to the GRCh38 reference genome.

### Data Analysis Tools

The scRNA-seq data were analyzed using the Seurat R package(Satija et al. 2015; Butler et al. 2018). Cell types were identified using singleR and the celldex::DatabaseImmuneCellExpression Data() function(Aran et al. 2019). K-means clustering was performed on NK cells based on gene expression using scikit-learn(Pedregosa et al. 2011). Outliers were defined as data points lying beyond two standard deviations from the mean of the distances. Uniform manifold approximation and projection (UMAP) (Becht et al. 2019) and t-distributed Stochastic Neighbor Embedding (t-SNE)(Cieslak et al. 2020) were applied to visualize the clusters. Plotting was done using matplotlib(Hunter 2007). Gene set enrichment analysis (GSEA) was done using the GSEA software 4.3.2(Subramanian et al. 2005a), applying the hallmark gene set collection(Liberzon et al. 2015) from the Molecular Signatures Database (MSigDB)(Subramanian et al. 2005b). A differential expression test was conducted for genes expressed in NK cells, excluding those used for clustering. The test compared gene expression between clusters within the sample, based on log-transformed counts, using a two-sample, two-tailed t-test. Considering the small sample size, our results focus on GSEA results with FDR-corrected significance, and DE test with significance replicated in all cases.

## Results

### Heterogeneous NK cell clusters with amplified changes of transcriptomic profiles in SV/HLHS

In our study, we observed that NK cell clusters tend to be heterogeneous (Fig.1). To test whether these heterogeneous NK cells are responsible for the DE genes, we examined the 1601 DE genes identified in NK cells from our previous study(Qu et al.). K-means clustering with these DE genes yielded high Silhouette scores with 2 clusters in all the 6 samples (Fig.2). As shown in Fig.3, the NK cells within the same sample can be divided into two distinct clusters. We then performed GSEA using the averaged difference between the two clusters across the three cases (Table 1, Supplementary Table 1). In one cluster, the gene sets HALLMARK_ANDROGEN_RESPONSE and HALLMARK_HYPOXIA are significantly upregulated, and HALLMARK_HEME_METABOLISM is significantly downregulated.

**Figure 1.**
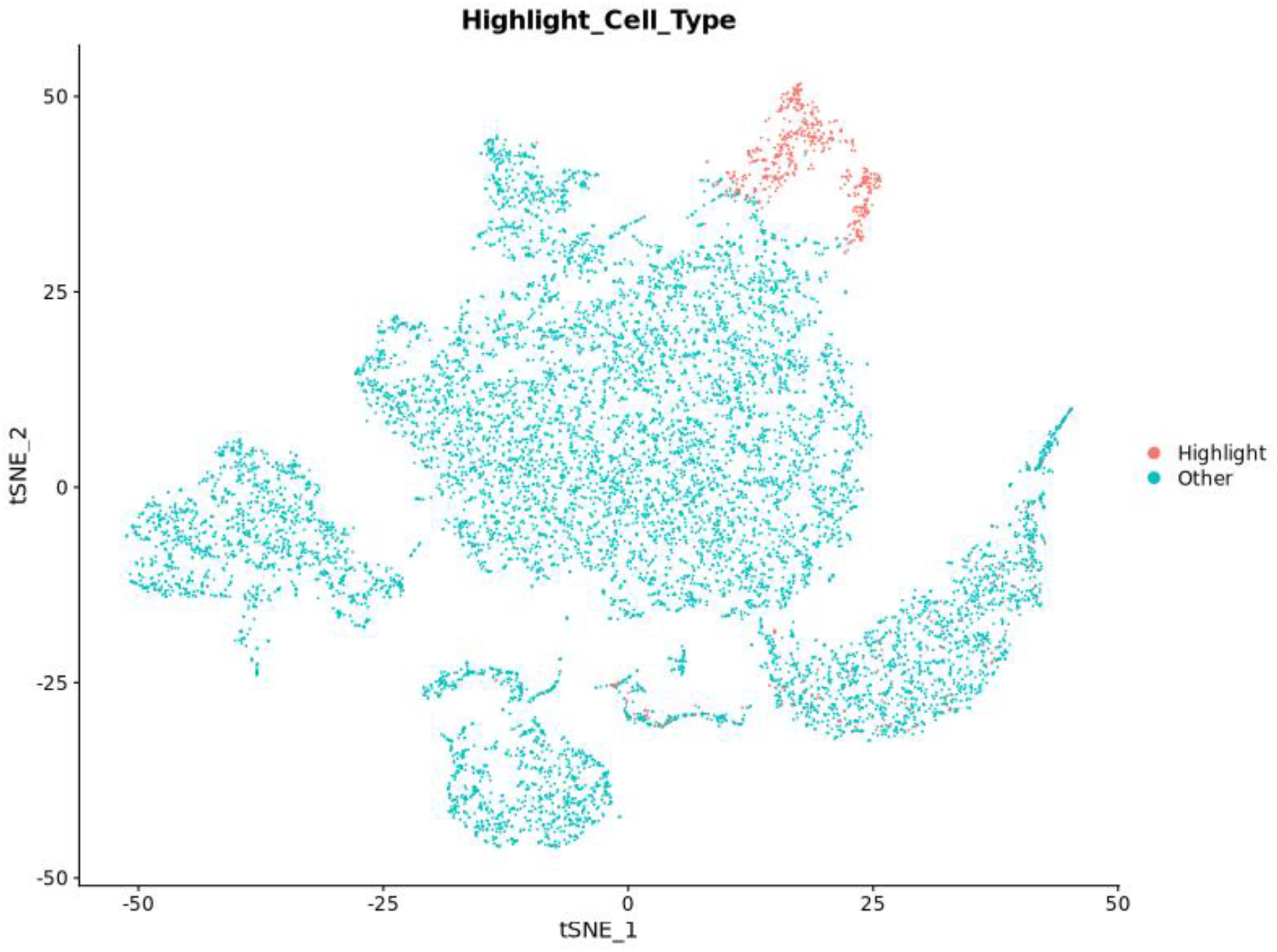
NK cell cluster versus other cell types visualized by t-SNE. NK cells are highlighted in red.

**Figure 2.**
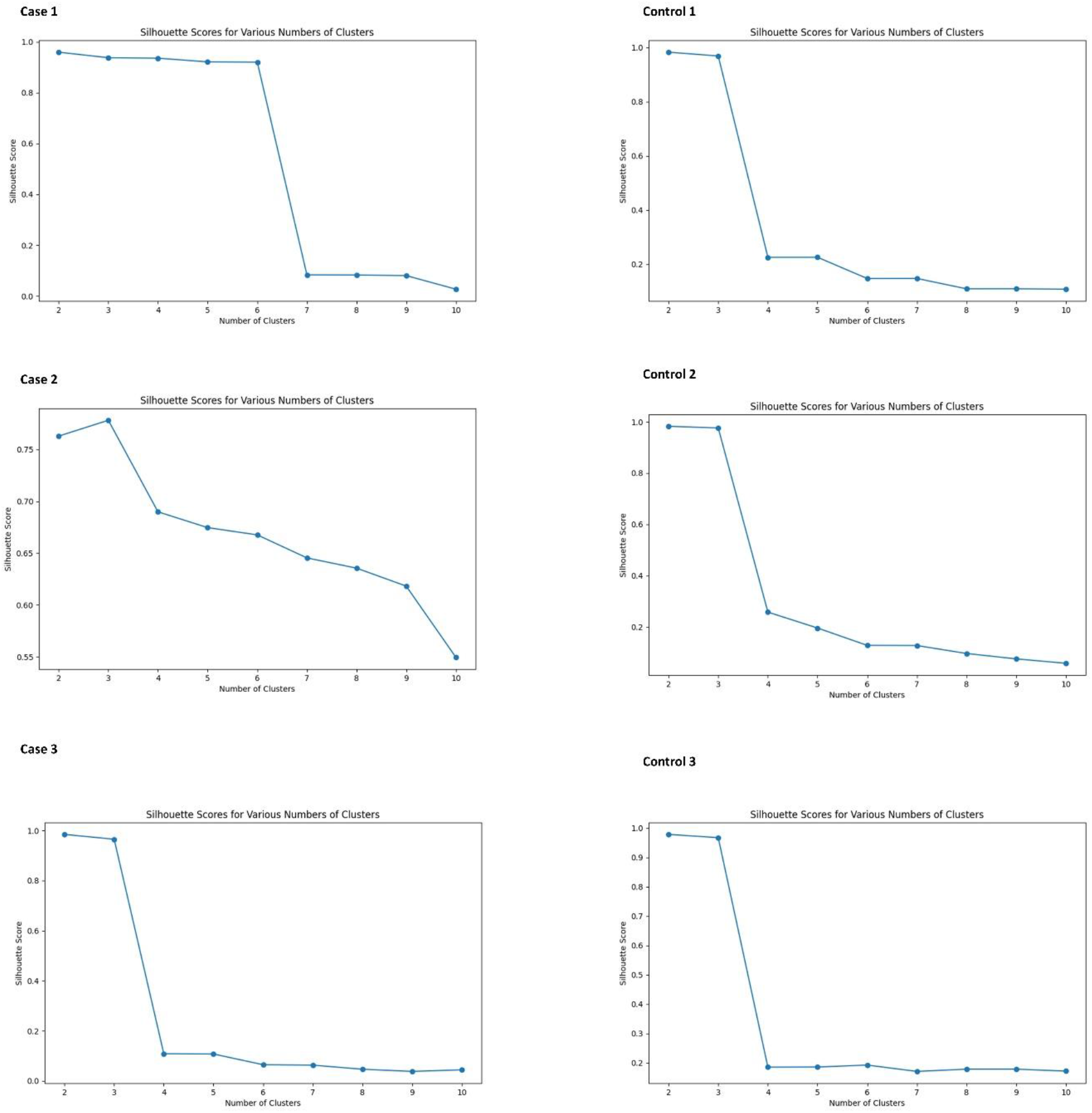
Silhouette scores for various numbers of clusters in K-means analysis of NK cells. All subjects show high Silhouette scores with two clusters.

**Figure 3.**
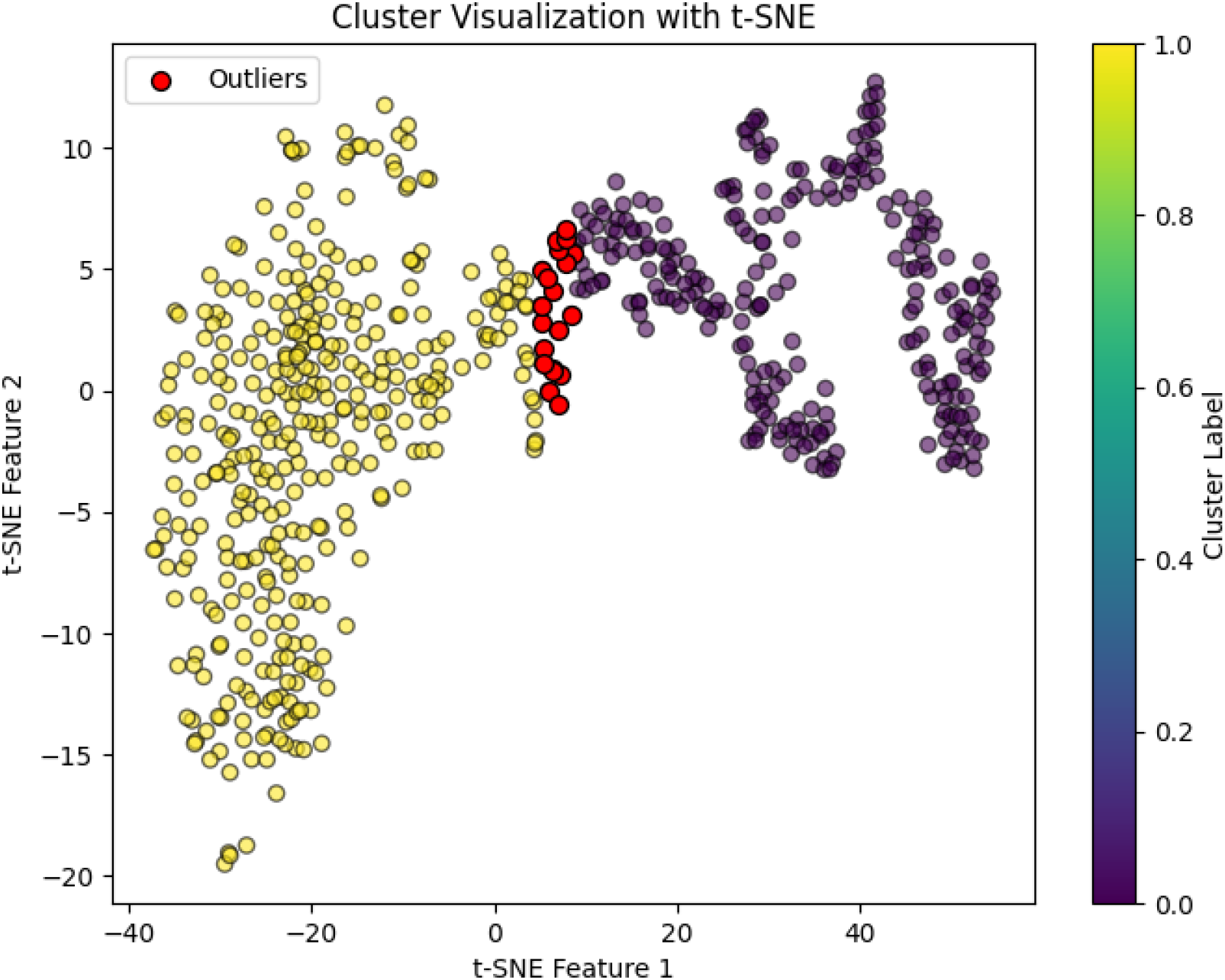
NK cell clusters visualized by t-SNE. Red dots indicate identified outliers.

**Table 1.** Gene sets with statistical significance identified by GSEA analysis.

| Gene set | SIZE | ES | NES | NOM p-val | FDR q-val |
| --- | --- | --- | --- | --- | --- |
| **1600 genes with differential expression in NK cells of SV/HLHS patient** |  |  |  |  |  |
| Cluster 1 vs Cluster 0: upregulated |  |  |  |  |  |
| HALLMARK_ANDROGEN_RESPONSE | 16 | 0.81 | 1.97 | 0 | 0 |
| HALLMARK_HYPOXIA | 20 | 0.66 | 1.64 | 0.009 | 0.041 |
| Cluster 1 vs Cluster 0: downregulated |  |  |  |  |  |
| HALLMARK_HEME_METABOLISM | 23 | -0.74 | -1.74 | 0 | 0.004 |
| Cluster expression in Cases vs Controls: upregulated |  |  |  |  |  |
| HALLMARK_ANDROGEN_RESPONSE | 16 | 0.81 | 1.94 | 0 | 0.004 |
| HALLMARK_OXIDATIVE_PHOSPHORYLATION | 52 | 0.63 | 1.8 | 0.001 | 0.021 |
| HALLMARK_INTERFERON_ALPHA_RESPONSE | 23 | 0.68 | 1.73 | 0.006 | 0.031 |
| HALLMARK_INTERFERON_GAMMA_RESPONSE | 38 | 0.63 | 1.71 | 0.004 | 0.027 |
| Cluster expression in Cases vs Controls: downregulated |  |  |  |  |  |
| HALLMARK_HEME_METABOLISM | 23 | -0.65 | -1.9 | 0.005 | 0.03 |
| **789 genes with differential expression between two clusters in NK cells** |  |  |  |  |  |
| Cluster 1 vs Cluster 0: upregulated |  |  |  |  |  |
| HALLMARK_ALLOGRAFT_REJECTION | 25 | 0.68 | 1.54 | 0 | 0.022 |
| Cluster 1 vs Cluster 0: downregulated |  |  |  |  |  |
| HALLMARK_UV_RESPONSE_DN | 15 | -0.43 | -1.58 | 0 | 0 |
| Cluster expression in Cases vs Controls: upregulated |  |  |  |  |  |
| Nonsignificant |  |  |  |  |  |
| Cluster expression in Cases vs Controls:<br>downregulated |  |  |  |  |  |
| HALLMARK_UV_RESPONSE_DN | 15 | -0.85 | -3.06 | 0 | 0 |

Compared to the controls, the upregulated androgen response and downregulated heme metabolism are more significant in the cases, as shown by Cluster expression in Cases vs Controls in Table 1. In addition, upregulation of the gene sets HALLMARK_OXIDATIVE_PHOSPHORYLATION, HALLMARK_INTERFERON_ALPHA_RESPONSE, and HALLMARK_INTERFERON_GAMMA_RESPONSE are more significant in Cluster 1.

### Other gene expression in the two clusters of NK cells

To further characterize the two NK cell clusters, we examined the expression of the two NK cell clusters for genes not used for the clustering. We identified 789 genes with DE between the two clusters in all three cases (Supplementary Table 2). GSEA analysis showed upregulated HALLMARK_ALLOGRAFT_REJECTION and downregulated HALLMARK_UV_RESPONSE_DN in Cluster 1 in cases. The downregulated HALLMARK_UV_RESPONSE_DN in Cluster 1 in cases is more significant than that in controls (Table 1).

## Discussion

In this study, we identified heterogeneous NK cell clusters with amplified changes of transcriptomic profiles in SV/HLHS. It is important to note that these gene expression changes in NK cells may not only arise as consequences of the heart condition but also potentially contribute to complications like viral infections or other cardiovascular issues.

### Distinct functional NK cell populations

In our analysis of NK cell populations, Cluster 1 exhibits distinct transcriptional profiles compared to Cluster 0 in term of the DE genes associated with SV/HLHS, marked by significant upregulation of gene sets including HALLMARK_ANDROGEN_RESPONSE and HALLMARK_HYPOXIA, alongside a notable downregulation of HALLMARK_HEME_METABOLISM. Androgen signaling may modulate NK cell immune functions, potentially tempering their cytotoxic activity while promoting survival and adaptation, particularly under stress(Liu et al. 2022). Concurrently, the increased expression of hypoxia-responsive genes likely enhances NK cells’ capability to operate in oxygen-deprived environments(Parodi et al. 2018). The reduced heme metabolism complements the other observed changes by potentially reducing oxidative stress and regulating metabolic responses(Dunaway et al. 2024), which may represent a shift towards energy-conserving metabolic processes. Collectively, these transcriptional changes suggest that NK cells in Cluster 1 are likely geared towards adaptation and survival in hypoxic or stress-related conditions. In contrast, Cluster 0 with different gene expression patterns might retain a higher cytotoxic capability and responsiveness, better suited for environments requiring rapid and robust immune reactions without the adaptive pressures of chronic stress.

### Distinct adaptations in Cluster 1 NK cells in SV/HLHS patients

The two clusters of NK cells observed in our study, present in both healthy controls and cases, likely represent distinct functional states tailored to specific immunological needs. Notably, marked differences in the expression profiles of these clusters between cases and controls were observed, as shown by Cluster expression in Cases vs Controls in Table 1. Cluster 1 in the cases shows significantly heightened androgen response and more pronounced downregulation of heme metabolism compared to controls. These changes may serve a dual role. This modulation could be protective, reducing potential tissue damage in chronic disease contexts such as seen in SV/HLHS. On the other hand, this reduced cytotoxicity might compromise the NK cells’ ability to clear pathogens effectively, potentially increasing the risk of infections. In addition, upregulation of the gene sets related to oxidative phosphorylation, and interferon responses, both alpha and gamma, are more significant in Cluster 1 in cases. Enhanced oxidative phosphorylation indicates that these NK cells have elevated energy production capabilities(Wang et al. 2020). Upregulated Interferon responses imply this cluster of NK cells is primed for responding to viral infections and potentially other pathogens(Gidlund et al. 1978). Altogether, upregulated HALLMARK_ANDROGEN_RESPONSE and downregulated HALLMARK_HEME_METABOLISM, plus these additional adaptations, suggest strategic modifications of NK cells to meet the demands of specific pathological states in prolonged exposure to pathogens or inflammatory conditions in SV/HLHS.

### Further characterization of the two NK cell clusters

In our extended analysis, we examined additional gene expressions within the two NK cell clusters beyond those initially used for clustering. This examination revealed 789 genes with DE between the two clusters across all cases. Specifically, Cluster 1 in cases exhibited upregulated HALLMARK_ALLOGRAFT_REJECTION and downregulated HALLMARK_UV_RESPONSE_DN. The upregulation of the allograft rejection gene set in Cluster 1 underscores a heightened immunological readiness to recognize and attack non-self cells, a crucial feature for defending against foreign tissues and potentially harmful pathogens(Hamada et al. 2021). The downregulation of genes associated with the UV response, including key regulators like *DYRK1A, ATXN1, ATP2B1, NIPBL, RUNX1*, and *SIPA1L1*, presents a complex scenario. These genes are crucial for various NK cell functions such as signaling, development, and effector responses(Voon et al. 2015; Matsuura et al. 2022). Their reduced expression might impair the NK cells’ capacity for regular immune surveillance and cellular migration. Furthermore, the downregulation of genes associated with the UV response in cases is significantly higher than controls. When considered together, these opposing trends in gene expression paint a picture of a dual nature in NK cells in Cluster 1, which could render these NK cells less effective in handling routine immune tasks, posing risks for viral infections and complicate the management of SV/HLHS.

Overall, these findings suggest that while NK cells in SV/HLHS are adapting to support survival in a challenging physiological environment, these adaptations may compromise their ability to manage additional stresses such as infections and inflammation. This nuanced view helps explain the dual role of these immune changes as both adaptations to and potential contributors to complications in SV/HLHS, underlining the need for targeted strategies to support immune function without exacerbating the underlying condition.

## Data Availability

All data produced in the present study are available upon reasonable request to the corresponding author.

## Ethics approval and consent to participate

All experimental protocols were approved by the Institutional Review Board (IRB) of the Children’s Hospital of Philadelphia (CHOP) with the IRB number: IRB 16-013278. Informed consent was obtained from all subjects. If subjects are under 18, consent was also obtained from a parent and/or legal guardian with assent from the child if 7 years or older.

## Funding

The study was supported by the Institutional Development Funds from the Children’s Hospital of Philadelphia to the Center for Applied Genomics, and The Children’s Hospital of Philadelphia Endowed Chair in Genomic Research to HH.

**Supplementary Table 1** Cluster expression of 1600 genes with differential expression in NK cells of SV/HLHS patient

**Supplementary Table 2** 789 genes with differential expression between two clusters in NK cells. *These genes were not used for clustering the NK cells. All of these genes showed differential expression with P<0.05 in each of the three cases.

